# Tracheal aspirate with closed suction device: a modified technique developed during COVID-19 outbreak

**DOI:** 10.1101/2020.08.24.20180802

**Authors:** Sofia Schverdfinger, Indalecio Carboni Bisso, Romina Famiglietti, Marcelo Di Grazia, Sabrina Di Stefano, Marcos Las Heras

## Abstract

**Background:** Bacterial superinfection as well as ventilation associated pneumonia (VAP) are both frequent events in critical care. During COVID-19 pandemic, usual diagnostic practices such as bronchoalveolar lavage and tracheal aspirate are limited due to their associated high risk of exposure for the operator. In order to set primary focus on the protection of health care personnel, a modified tracheal aspiration (M-TA) technique is developed and used for acquiring a lower respiratory tract of microbiological samples with a closed suction device.

**Methods:** Retrospective observational study to evaluate effectiveness of M-TA is conducted.

**Results:** A total of 33 M-TA samples were analyzed. In 66,6% of the cases, results led to a change in medical decision making. A 100% accuracy was achieved regarding COVID-19 diagnosis and a 56% bacterial growth-rate in cultives where VAP was suspected. No health care personnel have developed symptoms nor tested positive for COVID-19 during or after sample collection.

**Conclusion:** M-TA technique presented could be considered as a safe and effective procedure with low percentage of complications.

## INTRODUCTION

Coronavirus Disease 2019 (COVID-19) was defined as pandemic by the World Health Organization^1^ and affected more than 20 million people globally with confirmed cases in 215 countries. Since the beginning of the outbreak, the risk of viral transmission to healthcare personnel at the front line has been a global concern^2^ posing a challenge for healthcare systems in terms of managing human resources, supplies, and personal protective equipment. Therefore, it is key that all clinical decisions are based on prevention strategies to obtain the best value from the available resources^3^.

The collection of samples from the surface of the respiratory mucosa with nasopharyngeal swabs is a standard procedure used for diagnosis of COVID-19 in adults and children. Nevertheless, early data suggested relatively poor sensitivity of initial reverse transcription polymerase chain reaction (RT-PCR) tests from swabs^4^. False-negative results of nasopharyngeal swabs have direct implications for infection control and isolation rooms management. In addition to the previous, among critically ill patients the bacterial superinfection as well as a ventilator-associated pneumonia (VAP) are frequent events^5^, and usual practices such as bronchoalveolar lavage and traditional tracheal aspiration are limited due to their associated high risk of exposure for the operator^6,7^.

Setting primary focus on the protection of health care personnel, a modified tracheal aspiration (M-TA) technique is developed and used for acquiring a lower respiratory tract microbiological sample with a closed suction device. This technique proved to be useful not only in aiding COVID-19 diagnostic confirmation in patients with negative RT-PCR from swabs, but also in other respiratory infectious diseases when COVID-19 status remains unknown.

## METHODS

A retrospective analysis is conducted in medical records of patients with suspected or confirmed COVID-19 admitted to the intensive care unit (ICU) of a Level 3 University Hospital between June 1, 2020 and August 1, 2020. All patients included in the analysis are more than 18 years-old, underwent mechanical ventilation (MV), and required a lower respiratory tract microbiological sample for suspected bacterial superinfection or COVID-19 diagnosis. A description of epidemiological data, prior nasopharyngeal swab test-results and microbial rescue of M-TA from patients’ medical records is included.

### Study definitions

VAP suspicion was determined by the treating physician and was based on: new and persistent (48-h) or progressive radiographic infiltrate plus two of the following: temperature of 38°C or 36°C, blood leukocyte count of 10,000 cells/ml or 5,000 cells/ml, purulent tracheal secretions, and gas exchange degradation with increasing levels of positive end-expiratory pressure (PEEP) or fraction of inspired oxygen^8^. VAP was microbiologically defined, when the M-TA collected reached a count ≥10^5^ colony forming units per millilitre (UFC/ml). Only samples with less than 10 epithelial cells per field (100x magnification) were considered.

Suspected COVID-19 was determined following the Argentine National Ministry of Health definition^9^. COVID-19 diagnosis was microbiologically confirmed when RT-PCR from nasopharyngeal swabs or M-TA tested positive for SARS-CoV-2. All laboratory tests were processed in a College of American Pathologist-accredited laboratory.

Changes in medical decisions such as indication or withdrawal of isolation, initiation or suspension of antibiotics, and modifications in the antibiotic therapy were analyzed. Complications associated with the procedure were documented.

In order to perform tracheal aspiration, 2 operators donned with personal protective equipment (gown, cap, goggles, gloves, N-95 mask or equivalent, visor and second pair of gloves were required) entered the patient’s room where a high-efficiency particulate air filter was used. The collection of M-TA was performed using a 14 French siliconized polyvinylchloride (PVC) probe with a closed endotracheal suction system (Halyard Health®, United States) assembled with a sterile polypropylene collector bottle (CEEMED®, Argentina). A heat & moisture exchanger filter (HMEF) (Besmed®, Taiwan) was used to avoid virus particles dispersion, and a sterile T-63 tubing (BIOM®, India) was used for connecting pieces *(Figures 1 and 2)*. The PVC probe was introduced through the endotracheal tube or tracheostomy cannula until resistance was encountered (level of the carina in the trachea). This was followed by the release of the vacuum, and the probe was delicately removed using turning movements, until secretions were aspirated into the collector bottle. No saline solution was used for liquefy secretions, and strictly aseptic principles were followed. The M-TA technique performed is detailed in *Supplementary Material 1*.

**Figure 1.**
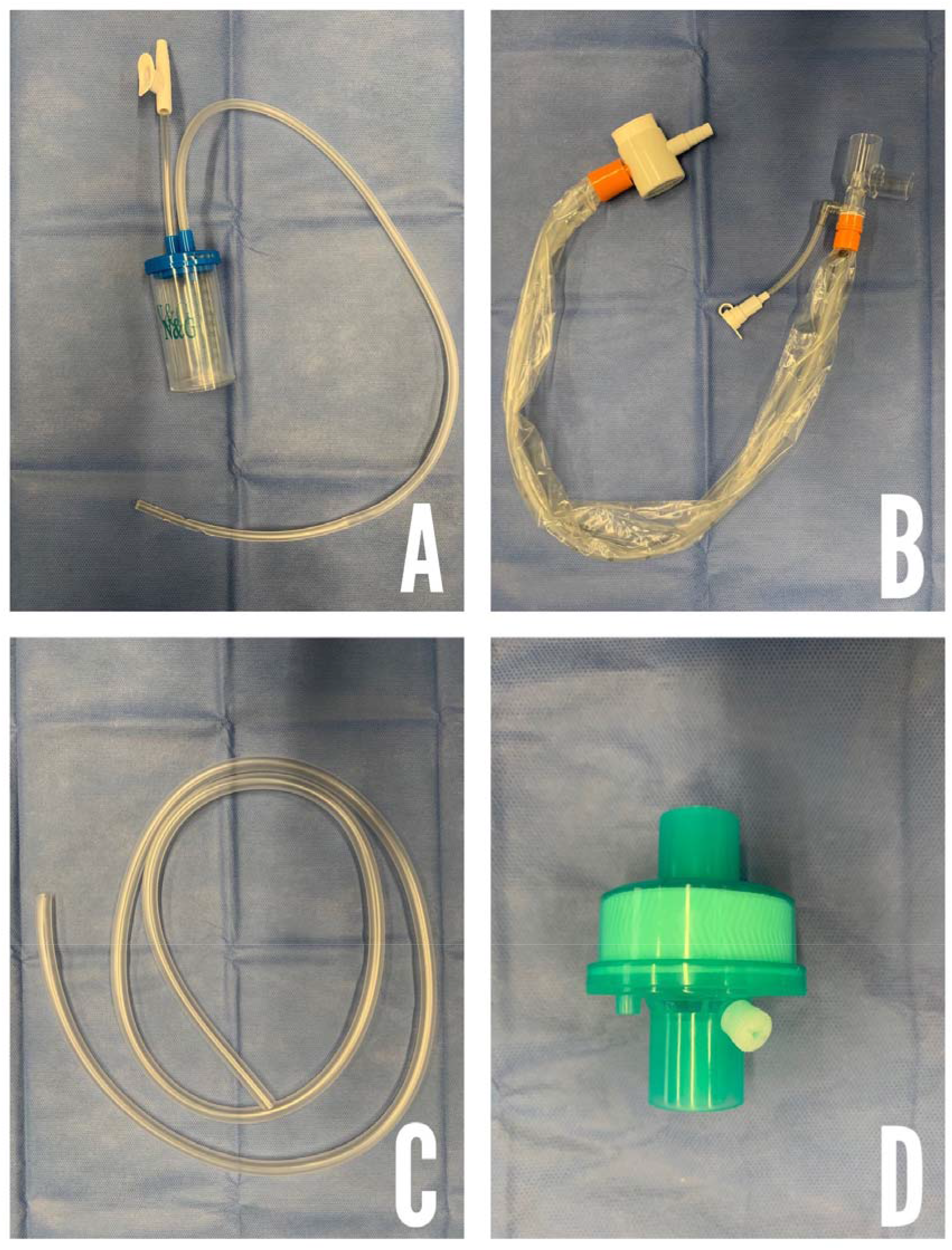
Required supplies. **A:** Collector bottle (also known as Lukens trap). **B:** Siliconized polyvinyl-chloride probe with a closed endotracheal suction system **C:** T63 tubing. **D:** Heat & moisture exchanger filter.

**Figure 2.**
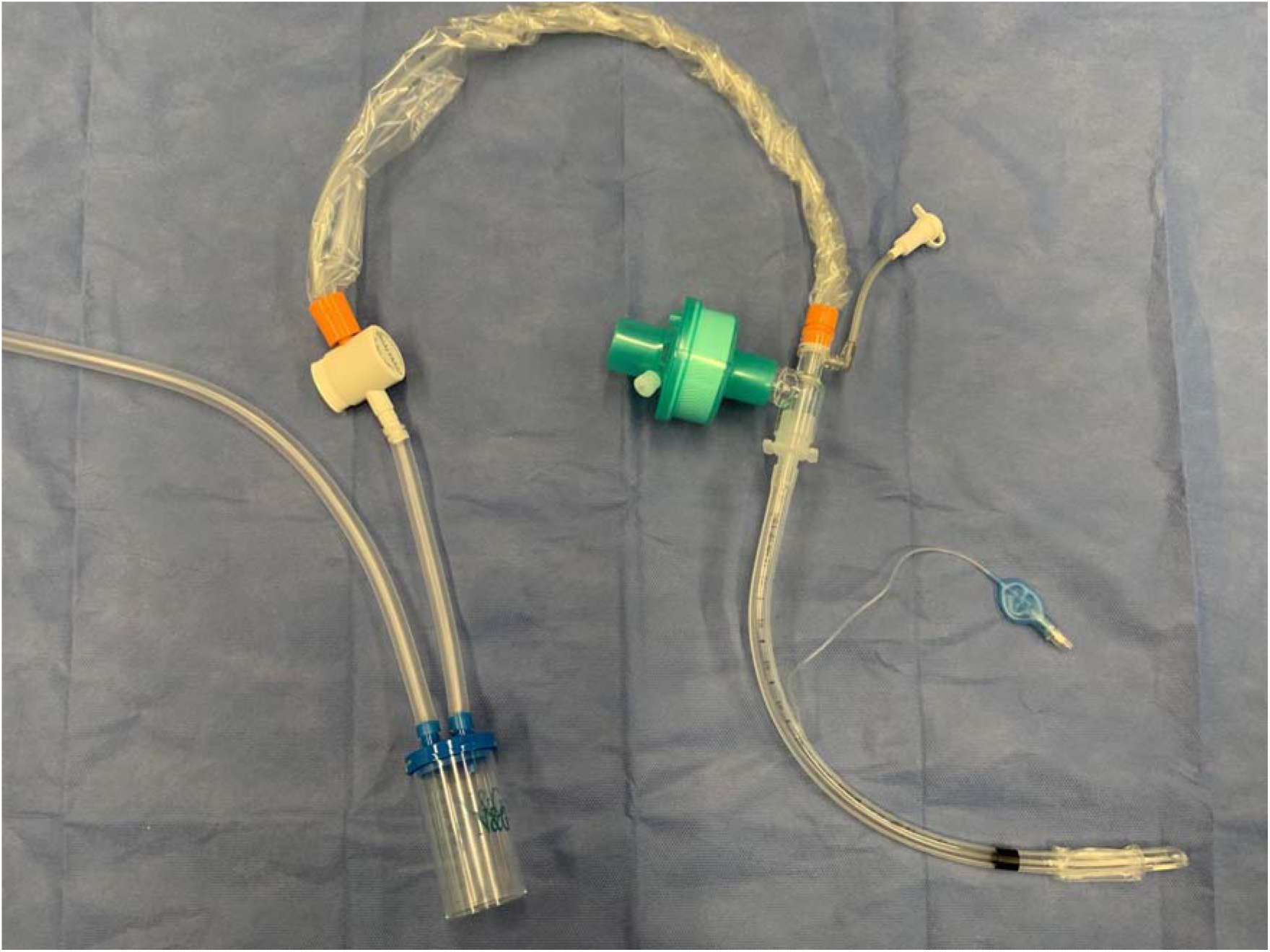
Closed suction device.

The data collection for this study was completed as part of the ICU follow-up clinic up to August 1st, 2020. This study was approved by the Ethics Committee of Hospital Italiano de Buenos Aires in June 2020, under protocol number 5678.

### Statistical analysis

Continuous variables were expressed as medians and interquartile ranges or simple ranges, as appropriate. Categorical variables were summarized as counts and percentages. No imputation was made for missing data. Given the fact that the cohort of patients in the study was not derived from random selection, all statistics are deemed to be descriptive only. RStudio developed by R-Tools Technology Inc was used for analysis.

## RESULTS

33 patients were included in the study; 10 were female (30,3%) and 23 men (69,6%). Median age was 71 years (interquartile range 64 - 78). Among the overall population, in 75,5% (25) of the cases, M-TA was performed due to VAP suspicion, and in 24,4% (8) of the cases M- TA was performed for COVID-19 diagnosis. Demographic and clinical characteristics of patients are shown in *Table 1*.

**Table 1.** Patient characteristics. Continuous variables are presented as median (and interquartile range), and categorical variables as count (%). APACHE II: Acute Physiology And Chronic Health Evaluation II.

|  |  |
| --- | --- |
|  | **n = 33** |
| Age, years | 71 (64 - 78) |
| Sex, male, % (no.) | 69,6% (23) |
| APACHE II score | 11 (9 - 14) |
| Charlson *score* | 5 (3 - 5) |
| Confirmed COVID-19 diagnosis prior to tracheal aspirate | 75,5% (25) |
| Mechanical ventilation duration, days | 16 (14 - 19) |
| ICU length of stay, days | 28 (23 - 32) |
| Hospital length of stay, days | 31 (23 - 46) |

**Table 2.**
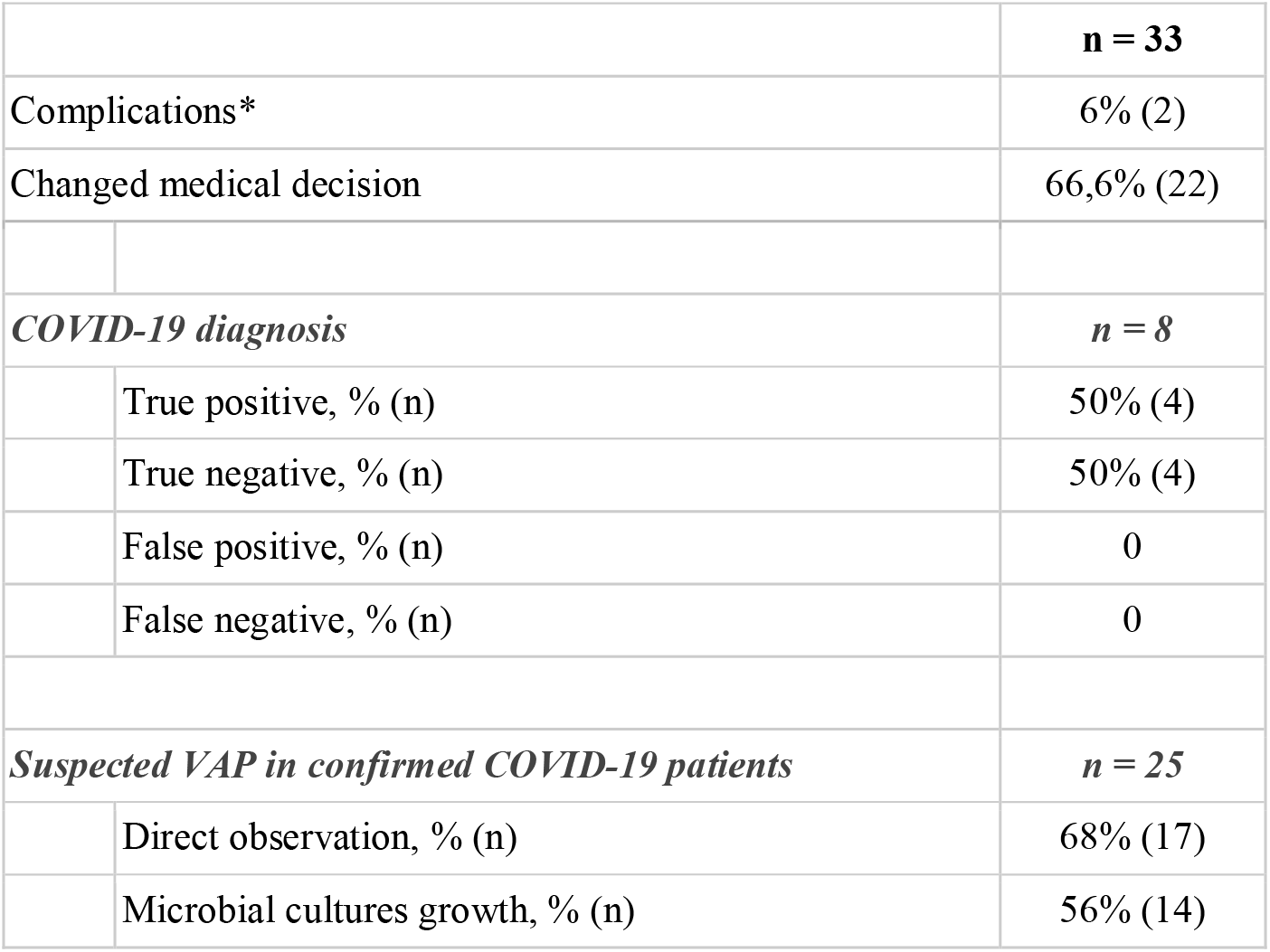
Results. Complications: hypotension due to propofol bolus (1), absence of tracheal secretions (1). Continuous variables are presented as median (and interquartile range), and categorical variables as count (%). VAP: Ventilator-associated pneumonia.

Of the 33 samples obtained, 22 (66,6%) led to a change in medical decision making. When technique was used for COVID-19 diagnosis, among a total of 8 patients 4 (50%) were a true positive and 4 (50%) true negative.

Analyzing microbiological samples due to suspected VAP in confirmed COVID-19 patients, 14 out of 25 samples (56%) presented microbial growth in cultives.

Among all 33 procedures performed, only 2 associated complications (6%) were observed: one hypotension episode due to propofol bolus, and one inability to obtain the sample due to absence of secretions.

## DISCUSSION

Incidence of negative RT-PCR results in SARS-CoV-2-positive patients is likely underreported. Due to the high risk of viral spread, an initial negative nasopharyngeal swab test should not alter clinical management in patients showing the constellation of symptoms consistent with COVID-19. The M-TA technique developed, achieved a 100% effectiveness rate regarding COVID-19 diagnosis. The true false reports were confirmed with SARS-CoV- 2 serologic studies so it is advised that when feasible, lower respiratory tract samples should be collected in order to help confirm the diagnosis^10^.

In the event that VAP was suspected, there was a high rate of germ rescue in the samples obtained. A higher revenue rate was observed in comparison with the one reported for bronchoalveolar lavage and conventional tracheal aspirate technique^12^. It is relevant to pinpoint that a key factor in decreasing mortality associated with VAP is the administration of adequate antibiotics as early as possible^11^.

Even though the size of the sample is small, the procedure described could be considered safe and effective with a low percentage of associated complications. Moreover, the technique developed involves minimal additional costs because it requires materials that are widely available.

Finally, it is worth to mention that none of the health care personnel involved have developed symptoms nor tested positive for COVID-19 during or after the data collection.

## Data Availability

All the information is available to be consulted.

## Data Availability Statement

all relevant data are within the manuscript and its Supporting Information files.

## Author Contributions

Dr. Schverdfinger had full access to all data in the study and takes responsibility for integrity of the data and accuracy of the data analysis.

## ACKNOWLEDGMENTS

The research team wants to specially thank Ignacio Fernández Ceballos for developing the technique. Also we would like to thank Maria de los Ángeles Magaz, Pablo Beber, Iván Huespe, Carolina Lockhart, Agustín Massó, Julieta González Anaya, Micaela Hornos, Marcela Ducrey, Pablo Coria, Germán Mayer and Juan Martín Nuñez Silveira for their valuable collaboration in this work.

